# The incubation period of 2019-nCoV from publicly reported confirmed cases: estimation and application

**DOI:** 10.1101/2020.02.02.20020016

**Authors:** Stephen A. Lauer, Kyra H. Grantz, Qifang Bi, Forrest K. Jones, Qulu Zheng, Hannah Meredith, Andrew S. Azman, Nicholas G Reich, Justin Lessler

**Author notes:** co-first authors (Stephen A. Lauer, Kyra H. Grantz).

## Abstract

A novel human coronavirus (2019-nCoV) was identified in China in December, 2019. There is limited support for many of its key epidemiologic features, including the incubation period, which has important implications for surveillance and control activities. Here, we use data from public reports of 101 confirmed cases in 38 provinces, regions, and countries outside of Wuhan (Hubei province, China) with identifiable exposure windows and known dates of symptom onset to estimate the incubation period of 2019-nCoV. We estimate the median incubation period of 2019-nCoV to be 5.2 days (95% CI: 4.4, 6.0), and 97.5% of those who develop symptoms will do so within 10.5 days (95% CI: 7.3, 15.3) of infection. These estimates imply that, under conservative assumptions, 64 out of every 10,000 cases will develop symptoms after 14 days of active monitoring or quarantine. Whether this risk is acceptable depends on the underlying risk of infection and consequences of missed cases. The estimates presented here can be used to inform policy in multiple contexts based on these judgments.

## Introduction

In December 2019, a cluster of severe pneumonia cases of unknown etiology was reported in Wuhan, Hubei province, China. The initial cluster was epidemiologically linked to a seafood wholesale market in Wuhan, though many of the initial 41 cases were later reported to have no known exposure to the market [1]. A novel strain of coronavirus belonging to the same family of viruses causing Severe Acute Respiratory Syndrome (SARS), Middle East Respiratory Syndrome (MERS), and the common cold was subsequently isolated from lower respiratory tract samples of four patients on 7 January 2020 [2]. Cases with no travel history to Wuhan have been reported, indicating likely human-to-human transmission [3]. On 30 January 2020, the World Health Organization declared that the 2019-nCoV outbreak constitutes a Public Health Emergency of International Concern [4]. Within 8.5 weeks of symptom onset of the first confirmed case (1 December 2019), there were more than 7,700 confirmed cases and 170 deaths reported worldwide [4]. On 31 January 2020, the United States Centers for Disease Control announced that all citizens returning from Hubei Province, China would be subject to mandatory quarantine for up to 14 days [5].

Currently, our understanding of the incubation period for 2019-nCoV is limited. An early analysis based on 34 confirmed cases in Chinese provinces outside Wuhan, using data on known travel to and from Wuhan to estimate the exposure interval, indicates a mean incubation period of 5.8 (95% CI: 4.6, 7.9) days, ranging from 1.3 to 11.3 days [6]. A more recent analysis based on 10 confirmed cases in China provided a similar estimate, showing a mean incubation period of 5.2 (95% CI: 4.1, 7.0) days [7]. This estimate is also in line with clinical reports of a familial cluster of 2019-nCoV in which symptom onset occurred 3-6 days after assumed exposure in Wuhan [1]. These estimates of the incubation period of 2019-nCoV are consistent with those of other known human coronaviruses, including SARS (mean 5 days, range 2-14 days;[8]), MERS (mean 5-7 days, range 2-14 days;[9]), and non-SARS human coronavirus (mean 3 days, range 2-5 days; [10]).

The incubation period can inform a number of important public health activities for infectious diseases, including active monitoring, surveillance, control, and modeling. Active monitoring requires potentially exposed individuals to contact local health authorities to report their health status every day. Understanding the length of active monitoring needed to limit the risk of missing 2019-nCoV infections is needed for health departments to effectively use limited resources. Here, we provide estimates of the incubation period of 2019-nCoV and the number of symptomatic infections missed under different active monitoring scenarios.

## Methods

### Data collection

We searched for news and public health reports of confirmed 2019-nCoV cases in areas with no known local transmission, including provinces, regions, and countries outside of Hubei. We searched for reports in both English and Chinese and abstracted the data necessary to estimate the incubation period of 2019-nCoV. Two authors independently reviewed the full text of each case report. Discrepancies were resolved by discussion and consensus.

For each case, we recorded the time of (i) possible exposure to 2019-nCoV; (ii) any symptom onset; (iii) onset of fever, and (iv) case detection. The exact timing of events was used when possible; otherwise, we defined upper and lower bounds for the possible interval of each event. For most cases, the interval of possible 2019-nCoV exposure was defined as the time between earliest possible arrival to Wuhan and latest possible departure from Wuhan. For cases without history of travel to Wuhan but with assumed exposure to an infectious individual, the interval of possible 2019-nCoV exposure was defined as the maximum possible interval of exposure to the infectious person (including time before the infectious individual was symptomatic). We assumed exposure always preceded symptom onset. If we were unable to determine the latest exposure time from the available case report, we defined the upper bound of the exposure interval to be the latest possible time of symptom onset. When earliest possible time of exposure could not be determined, we defined it as 1 December 2019, the date of symptom onset of the first known case [1]; we performed a sensitivity analysis for the selection of this universal lower bound. When the earliest possible time of symptom onset could not be determined, we assumed it to be the earliest time of possible exposure. When the latest time of possible symptom onset could not be determined, we assumed it to be the latest time of possible case detection. Data on individuals’ age, sex, country of residence, and possible exposure route were also collected.

### Statistical analysis

Cases were included in the analysis if we had information on the interval of exposure to 2019-nCoV and symptom onset. We estimated the incubation time using a previously described parametric accelerated failure time model [11]. For our primary analysis, we assumed that the incubation time follows a log-normal distribution, as seen in other acute respiratory viral infections [10]. We fit the model to all observations, and to subsets for only observations who experienced a fever and only observations detected inside or outside of mainland China as subset analyses. Finally, we also fit three other commonly used incubation period distributions (gamma, Weibull, and Erlang). We estimated median incubation time and a number of important quantiles (2.5%, 5%, 25%, 75%, 95%, and 97.5%) along with their bootstrapped confidence intervals for each model.

Using these estimates of the incubation period, we quantified the expected number of undetected symptomatic cases in an active monitoring program, adapting a method detailed by Reich *et al*. [12]. We account for varying durations of the active monitoring program (1-28 days) and individual risk of symptomatic infection (low: 1/10,000 chance that individual is infected; ‘medium’: 1/1,000; ‘high’: 1/100; ‘infected’: 1/1). For each bootstrapped set of parameter estimates from the log-normal model, we calculate the probability of a symptomatic infection developing after an active monitoring program of a given length for a given risk level. This model makes the conservative assumption that individuals are exposed to 2019-nCoV immediately preceding the active monitoring program and assumes perfect ascertainment of symptomatic cases that develop under active monitoring. We report the mean and 99th percentile of the expected number of undetected symptomatic cases for each active monitoring scenario.

All estimates are based on those who developed symptoms, and this work makes no inference about asymptomatic cases.

The analyses were conducted using the coarseDataTools and activemonitr packages in the R statistical programming language, version 3.5.3. All code and data are available at https://github.com/HopkinsIDD/ncov_incubation (release at time of submission at 10.5281/zenodo.3634050)[13].

## Results

We collected data from 101 confirmed cases of 2019-nCoV infection detected outside of the Hubei province before 28 January 2020 (Table 1). Of these, 34 (34%) were known to be female, 63 were male (62%), and 4 were of unknown sex. The median age was 52 years (IQR: 36.5-59). Cases were collected from 17 countries and regions outside mainland China (n=72) and 21 provinces within mainland China (n=29). The majority of cases (n=92) had a known recent history of travel to or residence in Wuhan; others had evidence of contact with travelers from Hubei or known cases. Amongst those who developed symptoms in the community, the median time from symptom onset to hospitalization was 2.7 days (range 0.2-26.2).

**Table 1.** Characteristics of confirmed 2019-nCoV cases.

|  |  |
| --- | --- |
|  | *n* = 101 |
| Age in years, median (IQR) | 52 (36.5, 59.0) |
| Sex, n (%) |  |
| Female | 34 (33.6) |
| Male | 63 (62.4) |
| Unknown | 4 (4.0) |
| Exposure to Wuhan, n (%) |  |
| Resident of Hubei province | 59 (58.4) |
| Known travel to Wuhan | 33 (32.7) |
| None/unknown | 9 (8.9) |
| Region of case detection (n): Mainland China (29), Singapore (10), Hong Kong (8), Taiwan (8), Thailand (8), Japan (5), Macau (5), United States (5), Australia (4), France (4), South Korea (4), Malaysia (4), Vietnam (2), Cambodia (1), Canada (1), Germany (1), Nepal (1), Sri Lanka (1) |  |

Fitting the log-normal model to all cases, we estimated the median incubation period of 2019-nCoV to be 5.2 days (95% CI: 4.4, 6.0). We estimated that fewer than 2.5% of infected individuals will display symptoms within 2.5 days (95% CI: 1.8, 3.6) of exposure, while symptom onset will occur within 10.5 days (95% CI: 7.3, 15.3) for 97.5% of infected individuals. The estimate of the dispersion parameter is 1.43 (95% CI: 1.22, 1.67). The estimated mean incubation period was 5.5 days.

To control for possible bias from symptoms of cough or sore throat, which could have developed from other more-common pathogens, we performed the same analysis on the subset of cases with known time of fever onset (n=61), using the time from exposure to onset of fever as the incubation time. Here, we estimated the median incubation period to fever onset to be 5.7 days (95% CI: 4.5, 7.0), with 2.5% of individuals experiencing fever within 2.9 days (95% CI: 2.2, 4.7) and 97.5% of individuals experiencing fever within 11.4 days (95% CI: 6.1, 17.8) of exposure.

Since assumptions about the occurrence of local transmission and therefore the period of possible exposure may be less firm within mainland China, we also performed an analysis of only cases detected outside mainland China (n=72). The median incubation period of these cases is 5.5 days (95% CI: 4,4, 7.0), with the 95% range spanning from 2.4 (95% CI: 1.7, 3.9) to 12.5 (95% CI 7.4, 19.0) days. Alternatively, cases who left mainland China may represent a subset of individuals with longer incubation periods, who were able to travel internationally before symptom onset within China, or may have chosen to delay reporting symptoms until they left China. Based on cases detected inside mainland China (n=29), the median incubation period is 4.6 days (95% CI: 3.4, 6.0), with a 95% range of 2.7 (95% CI: 1.2, 4.6) to 7.9 (95% CI: 3.9, 13.1) days.

We fit other commonly-used parameterizations of the incubation period: gamma, Weibull, and Erlang distributions. The incubation period estimates for these alternate parameterizations were similar to the log-normal model and to prior estimates. Compared to the log-normal, gamma, and Weibull estimates from Backer *et al* (2020), our estimates have similar medians but shorter tails, both on the upper and lower bounds of the incubation period (Figure 4).

Given these estimates of the incubation period, we predicted the number of symptomatic infections we would expect to miss over the course of an active monitoring program. We classified individuals as high-risk if they are assumed to have a 1 in 100 chance of developing a symptomatic infection after exposure. For an active monitoring program with a duration of 7 days, the expected number of symptomatic infections missed for every 10,000 high-risk individuals monitored is 19.7 (99th percentile: 39.4; Table 2). After 14 days, it is highly unlikely that further symptomatic infections would be undetected among high-risk individuals (mean: 0.6, 99th percentile: 4.5 undetected infections per 10,000 individuals). However, we note that there remains substantial uncertainty in the classification of individuals as being at ‘high’, ‘some’ or ‘low’ risk of being symptomatic, and that this method does not consider the role of asymptomatic infection.

**Table 2:** The expected number of symptomatic infections of 2019-nCoV that would go undetected during active monitoring given varying lengths of monitoring duration and risk of symptomatic infection after exposure. Estimates were generated from a probabilistic model using the incubation period estimates from the log-normal model.

| Mean estimated undetected symptomatic infections<br>per 10,000 monitored (99th percentile) |  |  |  |  |
| --- | --- | --- | --- | --- |
| Monitoring<br>duration | Low<br>(1 in 10,000) | Medium<br>(1 in 1,000) | High<br>(1 in 100) | Infected<br>(1 in 1) |
| 7 days | 0.2 (0.4) | 2.0 (3.9) | 19.7 (39.4) | 1,971.5 (3,940.2) |
| 14 days | 0.0 (0.0) | 0.1 (0.4) | 0.6 (4.5) | 64.4 (449.6) |
| 21 days | 0.0 (0.0) | 0.0 (0.1) | 0.1 (0.8) | 5.3 (75.3) |
| 28 days | 0.0 (0.0) | 0.0 (0.0) | 0.0 (0.2) | 0.8 (17.0) |

## Discussion

Here, we present estimates of the incubation period for the novel coronavirus (2019-nCoV) that emerged in Wuhan, Hubei Province, China. We estimate the median incubation period of 2019-nCoV to be 5.2 days and expect nearly all infected individuals who experience symptoms will do so within 11 days of infection. We find that the current period of active monitoring recommended by the US CDC of 14 days is well supported by the evidence [5]. Symptomatic disease is frequently associated with transmissibility of a pathogen. However, given recent evidence of asymptomatic transmission of 2019-nCoV [3][14], we note that time from exposure to onset of infectiousness (latent period) may be shorter than the incubation period estimated here.

While our results support current proposals for the length of quarantine or active monitoring of those potentially exposed to 2019-nCoV, in extreme cases longer monitoring periods might be justified. Among those who are infected and will go on to develop symptoms, we expect 64 in 10,000 will do so after the end of a 14 day monitoring period (Table 2), and our estimates do not preclude this estimate being higher. Hence, in instances where there is a high probability of an infectious exposure (e.g., a healthcare worker who cared for an 2019-nCoV patient while not wearing personal protective equipment), it may be prudent to consider symptoms after this period as potentially 2019-nCoV associated, and potentially extend the period of active monitoring.

There are several important limitations to this analysis. Our data include early case reports, with some associated uncertainty in the intervals of exposure and symptom onset. We have attempted here to use conservative bounds of possible exposure and symptom onset where exact times were not known, but there may be further inaccuracy in this data that we do not consider here. We have exclusively considered reported, confirmed cases of 2019-nCoV, which may overrepresent hospitalized individuals and others with severe symptoms. The incubation period for these severe cases may differ from that of less severe or subclinical infections, and is not typically an applicable measure for those with asymptomatic infections.

Our model assumes a constant risk of 2019-nCoV infection in Wuhan from 1 December 2019 to present, based on the date of symptom onset of the first known case. This is a simplification of infection risk, as the outbreak has shifted from a likely common-source outbreak associated with a seafood market to human-to-human transmission. Moreover, phylogenetic analysis of 38 2019-nCoV genomes suggests that the virus may have been circulating prior to December [15]. To test the sensitivity of our estimates to that assumption, we performed an analysis where cases with unknown lower bounds on exposure were set to 1 December 2018, a full year earlier than in our primary analysis. Changing this assumption had little effect on the estimates of the median (<0.1 days different), and only slightly increased our estimates of the 97.5th quantile of the incubation period (0.4 days longer than for the overall estimate).

This work provides additional evidence for a median incubation period for 2019-nCoV of approximately five days, similar to SARS. Assuming infection occurs at the initiation of monitoring, our estimates suggest that 64 out of every 10,000 cases will develop symptoms after 14 days of active monitoring or quarantine. Whether this rate is acceptable depends on the expected risk of infection in the population being monitored, and considered judgment about the cost of missing cases [12]. Combining these judgements with the estimates presented here can help public health officials to set rational and evidence-based 2019-nCoV control policies.

## Data Availability

All code and data for this manuscript is available on github.

https://doi.org/10.5281/zenodo.3634049

## Acknowledgements

We are very grateful to all who have collected, prepared, and shared data throughout this outbreak. We are particularly grateful to Dr. Kaiyuan Sun and Dr. Moritz Kraemer for sharing their data on confirmed infections within and outside of Wuhan and the Johns Hopkins Center for Systems Science and Engineering.

---

**Figure 1.**
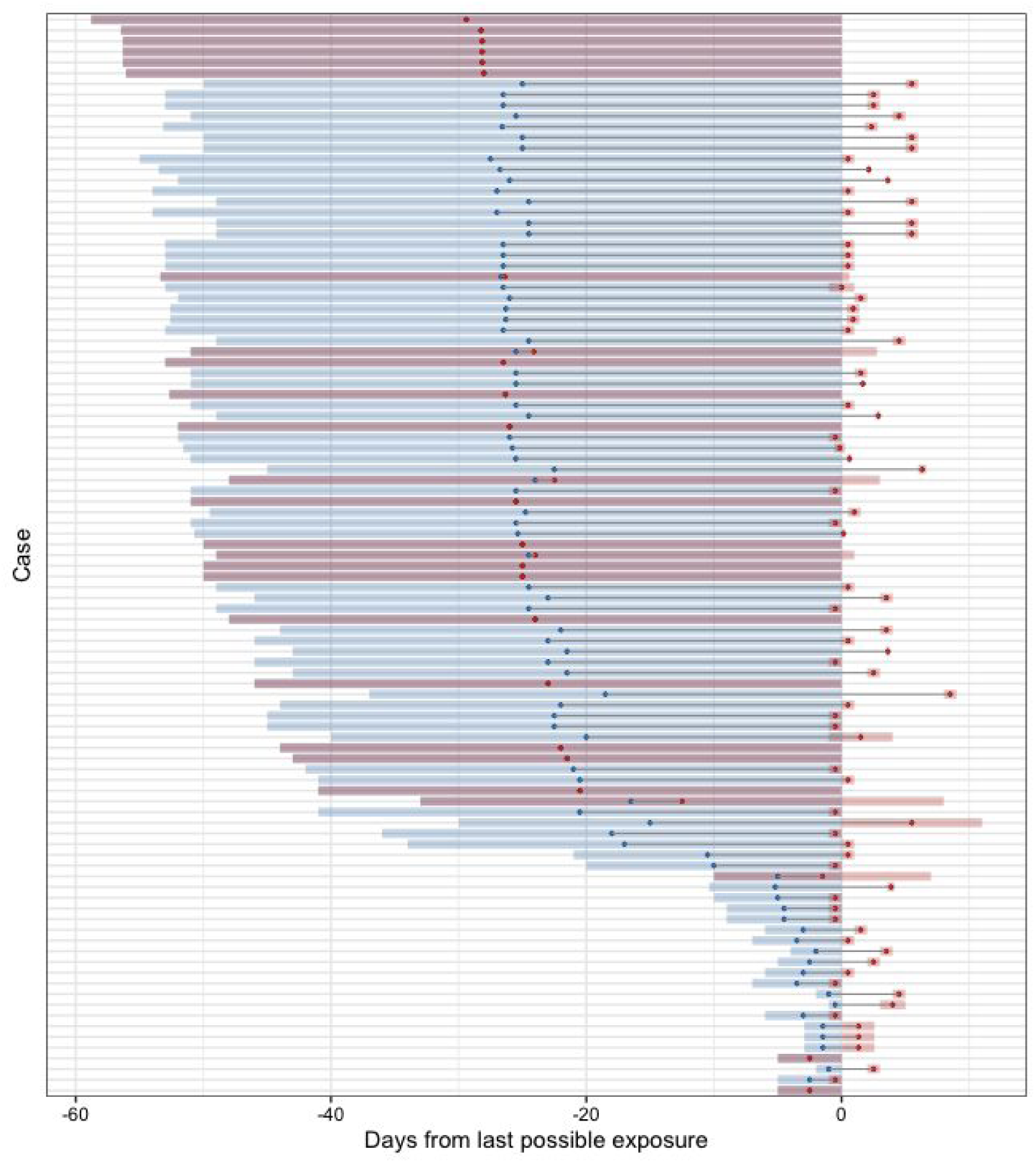
Novel coronavirus (2019-nCoV) exposure (blue) and symptom onset (red) times for 101 confirmed cases. Shaded regions represent the full possible interval of exposure and of symptom onset; points represent the midpoint of these intervals.

**Figure 2:**
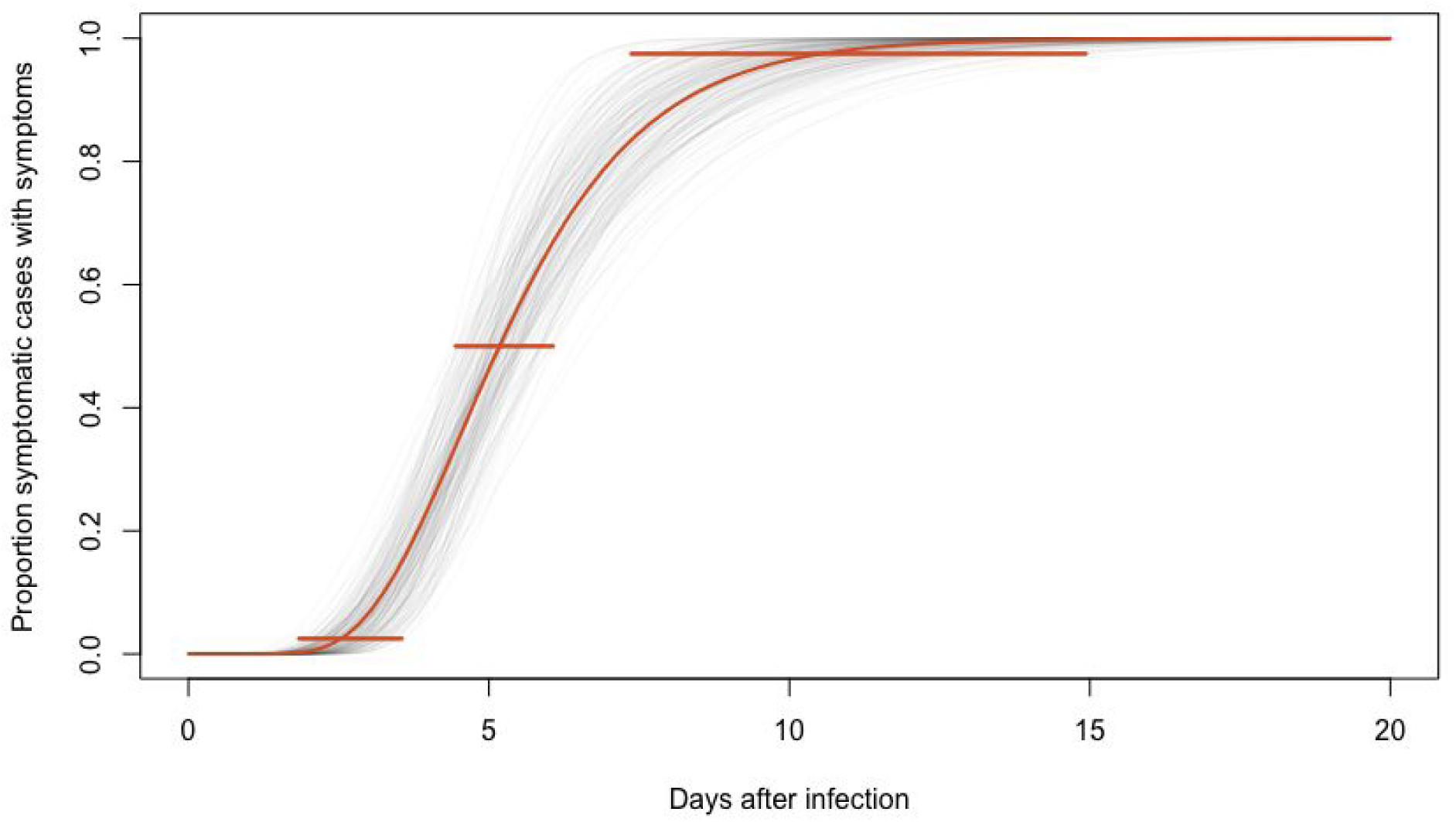
Cumulative distribution function of the novel coronavirus (2019-nCoV) incubation period estimate from the log-normal model. The estimated median incubation period of 2019-nCoV was 5.2 days (95% CI: 4.4, 6.0). We estimated that fewer than 2.5% of infected individuals will display symptoms within 2.5 days (95% CI: 1.8, 3.6) of exposure, while symptom onset will occur within 10.5 days (95% CI: 7.3, 15.3) for 97.5% of infected individuals. Horizontal bars represent the 95% confidence intervals of the 2.5, 50, and 97.5 percentiles of the incubation period distribution. The estimate of the dispersion parameter is 1.43 days (95% CI: 1.22, 1.67).

**Figure 3:**
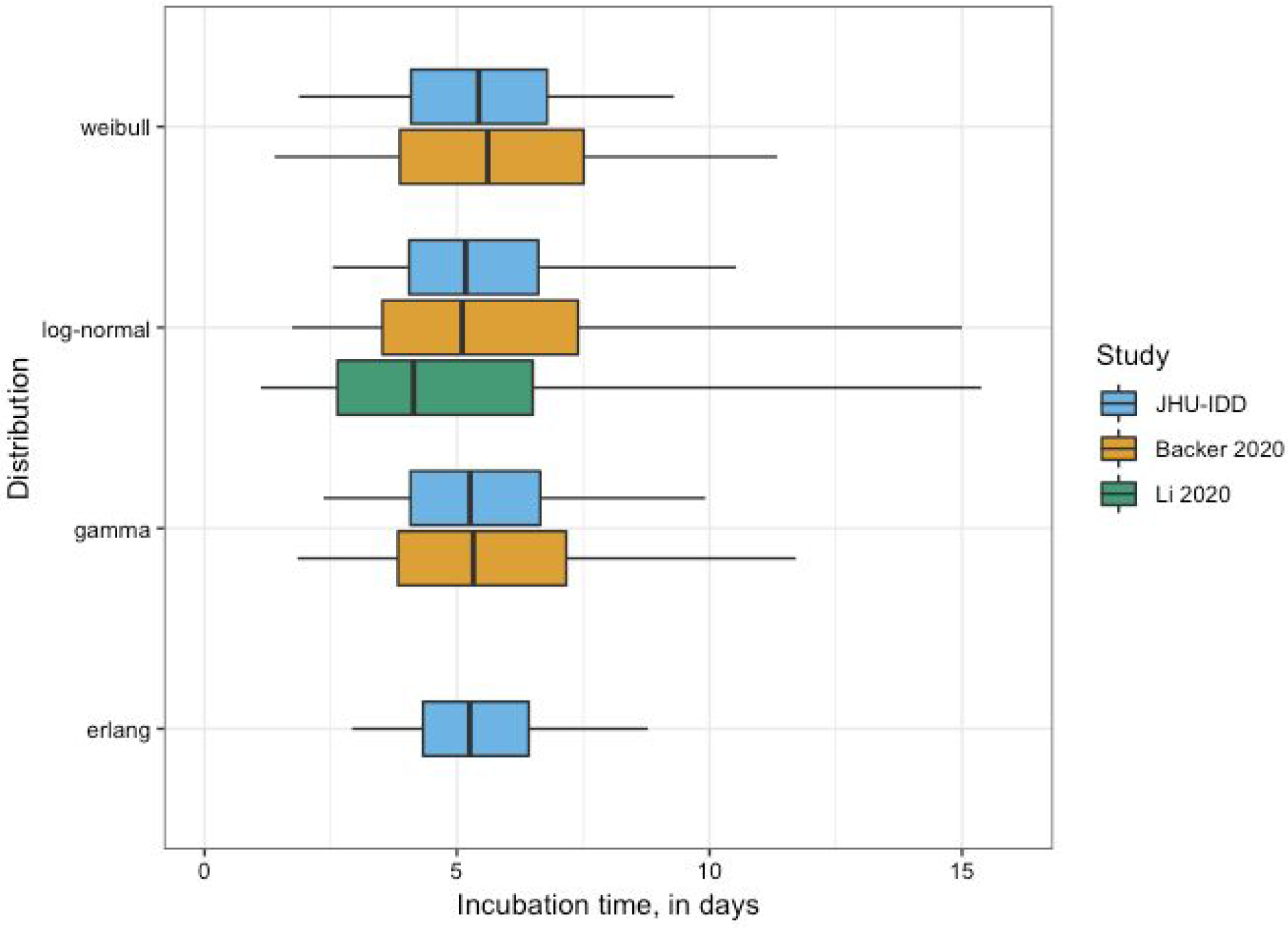
Comparison of estimates for median incubation period of novel coronavirus (2019-nCoV). In our analysis, the gamma distribution has an estimated shape parameter of 7.92 (95% CI: 3.97, 24.98) and a scale parameter of 0.69 (95% CI: 0.21, 1.52); the Weibull distribution has an estimated shape parameter of 3.11 (95% CI: 2.2-6.08) and a scale parameter of 6.11 (95% CI: 5.19, 7.25); and the Erlang distribution has an estimated shape parameter of 14 (95% CI: 5, 21) and a scale parameter of 0.4 (95% CI: 0.26, 1.11).

